# A digital app-supported general practitioner-led weight loss program improves food literacy and intake in adults in rural Germany: Secondary outcomes of the HAPpEN pragmatic trial

**DOI:** 10.1101/2025.08.21.25334205

**Authors:** Anna Weber, Marika Haderer, Reiner Hofmann, Mirna Al Masri, Natascha von Schau, Laura M. König

## Abstract

Obesity prevalence is rising globally and especially in rural areas. Interventions promoting dietary behavior change specifically in rural populations are urgently needed to address this issue. This analysis used data from HAPpEN, a multicomponent, general practitioner (GP)-led and digitally supported obesity management intervention specifically developed to support weight loss in rural adults. It tested whether (1) the intervention induced changes in food literacy and intake and their relationships to changes in body weight, (2) these effects were modulated by the preferred decision-making style, and (3) the intervention induced eating disorder symptoms. Only intervention completers (*N* = 61) were included in the analysis.

Outcomes were assessed at baseline, six months into the program, and immediately after completing the one-year program. Several aspects of food literacy including daily food planning improved significantly after six months, although the total food literacy score only improved significantly after one year. Food intake improved significantly after six months and stayed stable thereafter. At six months, changes in food intake were also related to changes in body weight. Intervention effectiveness may have dependent on the preferred decision-making style. Eating disorder symptomatology was not worsened through the program.

Successful weight loss induced by HAPpEN is likely driven by changes in food literacy and intake. Offering a range of target foods, graded tasks and several complementary intervention components allows for tailoring to varying patient needs, levels of knowledge and behavior, and therefore increases the likelihood for intervention success. This trial was registered at the German Clinical Trial Register (DRKS00033916).

## 1. Introduction

The prevalence of obesity is rising globally and forecasted to affect more than half of the global adult population by 2050 [1]. Action is urgently required to promote healthier lifestyles and to prevent detrimental effects on individual health and well-being [2] as well as on healthcare systems and economies [3, 4]. Obesity prevalence is even higher in rural vs. urban areas [5], which also translates into higher prevalence of noncommunicable diseases such as Diabetes mellitus type 2 and coronary heart disease [6]. This discrepancy is partly due to physical barriers to engage in healthier behaviors [7]. For example, access to supermarkets and food outlets that primarily sell unprocessed foods is restricted in rural areas [8] resulting in unhealthier eating patterns in rural populations [9]. Support to promote healthier food intake in these populations is thus crucial.

Especially in more remote locations, access to healthcare practitioners may be limited. It is assumed that digital technology, including websites and smartphone apps, may close this gap in care provision, also in the context of obesity prevention and management [10]. Indeed, digital interventions are promising (dietary) behavior change tools [11]. Yet, evaluations of weight management programs in general and digitally enhanced programs specifically in rural areas remain sparse [12], especially outside the United States [13]. This is potentially due to universities and study centers being located in urban areas, which increases travel distances for rural populations [14]. Transdisciplinary collaboration may close this gap, for instance by involving local general practitioners (GPs) as points of contact and support in recruitment and taking relevant measurements [15].

To address the need for obesity prevention and management in rural areas, the HAPpEN (“Hausarzt-zentriertes Adipositas-Präventionsprogramm: Exercise & Nutrition“) pragmatic trial was conducted in rural Germany and in close collaboration between a university campus in a rural area and local GPs [16]. It tested a multicomponent, GP-led intervention with digital app support to explore innovative, scalable strategies for obesity management; the intervention significantly reduced the patients’ body weight and improved several metabolic risk factors throughout the one-year program, with changes being most pronounced in the first six months [17]. Given that obesity often results from energy imbalance (amongst others due to processed carbohydrates) [18], promoting healthier dietary behaviors is a core component of obesity management. It is thus to be expected that changes in food intake occurred that drove reductions in body weight. An important prerequisite of dietary behavior change, in turn, is food and nutrition literacy [19], i.e. “people’s knowledge of healthy eating and their capability to purchase and prepare healthy foods” (Poelman et al. [20], p. 1). Interventions thus may need to target food literacy to induce dietary behavior change.

The present analysis thus evaluated food intake and literacy as secondary outcomes of the HAPpEN trial. First, this analysis aimed to test the effects of the program on food intake and literacy over the course of the one-year program to provide insights into underlying mechanisms of the intervention’s effects on body weight. It was assumed that food literacy in intake would improve significantly throughout the program, which was tested by comparing food literacy and intake between baseline, at six months into the program, and immediately after completing the one-year program. Second, it aimed to test whether the program was equally effective for participants with a preference for deliberation and intuition in eating decision-making. Prior research suggests that especially digital weight management programs may employ behavior change techniques that primarily cater to the needs of individuals with a strong preference for deliberation, i.e. planning of food intake [21]. This may limit the effectiveness of interventions for individuals with a stronger preference for intuition, i.e. those who prefer to rely on their gut feeling; the present analysis thus provides insights into a potential need for tailoring weight management interventions according to the preferred decision-making style. Third, it aimed to test for potential negative effects of the intervention by investigating changes in eating disorder symptomatology. It has been hypothesized that self-monitoring interventions in particular may promote eating disorders due to their restrictive focus [22]; this study therefore tested this assumption by comparing symptomatology before and after the intervention.

## 2. Methods and materials

### 2.1. Study design and participants

The HAPpEN single-group pragmatic trial evaluated a GP-centered, digital app-supported obesity management program focusing on exercise and nutrition in a rural setting [16]. Six GPs acted as primary points of contact for their patients, who were enrolled in the program for one year.

The participants were recruited in GP practices between May and June 2023. People between 18 and 65 years with a body mass index (BMI) ≥ 30 kg/m^2^ and the ability and willingness to use a digital, app-based self-monitoring diary were eligible to participate. Reasons for exclusion as well as detailed information on participants and recruitment can be found elsewhere [16, 17]. Since this was a pragmatic trial conducted in a primary care setting and participants were recruited via the six participating GP practices, no formal sample size calculation was conducted; we aimed to recruit 100 participants since this was deemed feasible to handle, given the resources available for the trial.

The HAPpEN trial has been approved by the University of Bayreuth ethics committee (Az. O 1305/1 – GB) and was retrospectively registered at the German Clinical Trial Register (DRKS00033916). All participants provided written informed consent prior to the start of the study.

### 2.2. Intervention

See the study protocol [16] and main outcome paper [17] for a detailed description of the intervention. In brief, in the HAPpEN obesity management program, GPs served as the central coordinators and supporters of their patients’ weight loss efforts. Based on an initial assessment, they developed individual therapy plans that focused on nutrition and physical activity. In monthly appointments, the participants discussed their progress and challenges while receiving ongoing motivational support. The program also included introductory group sessions that provided information on weight loss, dietary and physical activity guidelines; physiotherapy sessions to support increased physical activity; and a range of group activities for social support. Additionally, participants were introduced to the HAPpEN app, which was available as a smartphone or web application and provided access to self-monitoring tools for goal progress, setting graded tasks, and knowledge and instruction via a series of modules for self-paced learning. An overview and description of the included behavior change techniques is available in the study protocol [16].

Several components of the intervention were specifically designed to promote food literacy and foster healthier food intake:

(1) Immediately after enrolment, participants took part in a 90-minute group session on dietary recommendations, with a specific focus on reducing energy density of the consumed foods and drinks [23]. This session also introduced the concept of graded tasks to facilitate goal setting. The core areas of dietary behavior and food literacy targeted are listed in Table 1. Enrolled participants also took part in another 90-minute group session on behavior change strategies, where they were introduced to relevant techniques such as goal setting.
(2) For each core area of behavior change, a stepwise framework was developed to derive graded tasks, which were used to tailor the intervention to individual needs and circumstances and to allow to set challenging yet achievable goals [24]. Each month, participants were asked to choose two to three topics to work on from a list that also included several physical activity-related topics. For each chosen topic, participants were asked to select a challenging yet achievable goal out of a list of five to eight options ranging from “I am not aware of how to address this topic in my daily life” (aiming at self-monitoring their behavior to identify areas for change) to “reduce unhealthy habits and improve lifestyle on up to six to seven days/week” (see [16, 17] for the exact wording and tasks for all behaviors).
(3) Participants were encouraged to use the digital app to self-monitor goal progress.
(4) Several cooking events with the study staff were offered on a voluntary basis.

**Table 1.** Dietary behaviors and food literacy topics addressed in the HAPpEN intervention.

| Dietary behavior | Food literacy |
| --- | --- |
| - to replace sugary soft drinks and alcohol consumption with energy-free drinks such as water and unsweetened tea<br>- to reduce high-energy snacks such as crisps, sweets etc. and establish healthy snack alternatives<br>- to stop consuming fast food and ready-made products with high energy density<br>- to consume five portions of fruits and vegetables a day<br>- to increase the daily fiber content by replacing white flour with wholegrain products or implementing pulses<br>- to use different protein sources, limiting meat consumption to 1-2 per week if necessary<br>- to reduce the fat content of their diet | - to develop a healthy eating rhythm with three main meals<br>- to establish a healthy breakfast<br>- to focus on a healthy main meal<br>- to avoid snacks between meals in the form of a second breakfast, cake with coffee in the afternoon, snacks when coming home after work or before going to bed.<br>- to avoid eating late in the evening and to have the last meal at least three hours before sleeping<br>- to exercise a slow and conscious way of eating with thorough chewing and avoiding distractions such as mobile phones, televisions, newspapers etc. |

### 2.3. Measures

A range of physiological, psychological and behavioral measures were taken at baseline (T0), six months after enrolling in the program (T6) and at the end of the one-year program (T12). The full list of measures is presented in the study protocol [16]. In the following, only the measures relevant to the presented analysis are described.

#### 2.3.1. Outcome measures

Food intake was assessed with a modified Food-Frequency Questionnaire (FFQ) [25, 26]. Participants indicated consumption frequency on a six-point scale (1 = almost daily to 7 = never) for a comprehensive selection of foods relevant to weight gain and weight loss. A healthy eating index (HEI) was calculated based on 15 food groups [26], based on which individuals’ food consumption was classified into a favorable dietary pattern (scores of 16 and above), a normal dietary pattern (scores of 14 and 15), and an unfavorable dietary pattern (scores of 13 and lower). In addition to the score, changes in the consumption frequency of the individual food groups were also evaluated. For this, the coding was inversed so that higher values indicate more frequent consumption. Additionally, a comprehensive assessment of beverage consumption was developed with seven frequency categories (original coding: 1 = more than five times a day to 7 = rarely/never; inverse coding: 1 = rarely/never to 7 = more than five times a day) based on the German DEGS study [27]. Food intake was assessed at T0, T6 and T12.

Food literacy was assessed with the German version [28] of the Self-Perceived Food Literacy Scale (SPFLS) [20]. It includes 29 items to assess knowledge, skills, and confidence related to food and nutrition which are grouped into eight domains: food preparation skills (six items), resilience and resistance (six items), healthy snack styles (four items), social and conscious eating (three items), examining food labels (two items), daily food planning (two items), healthy budgeting (two items), and healthy food stockpiling (four items). Responses were recorded on a five-point Likert scale (1= not at all/never to 5 = yes/always), with negative items reversed, and the mean calculated per subscale and for the total scale, so that a high score indicates high food literacy. For the subscales”food preparation skills”,”social and conscious eating” and”resilience and resistance”, certain items reduced the reliability of their subscale and thus were removed. Thus, in the presented analysis”food preparation skills” consist of five items α_T0_ = 0.79 (vs. six items, α _T0_ = 0.53; item removed:” Do you find it difficult to prepare a meal with more than five fresh ingredients?”),”social and conscious eating” of two items α _T0_ = 0.66 (vs. three items, α _T0_ = 0.25; item removed”Do you engage in any other activities while eating?”) and”resilience and resistance” with five items α _T0_ = 0.65 (vs. six items, α _T0_ = 0.48; item removed:”Do you eat the total contents of a bag or container of crisps, candies or cookies in one go?”). See Table 2 for all reliability indices. Food literacy was assessed at T0, T6 and T12.

**Table 2.**
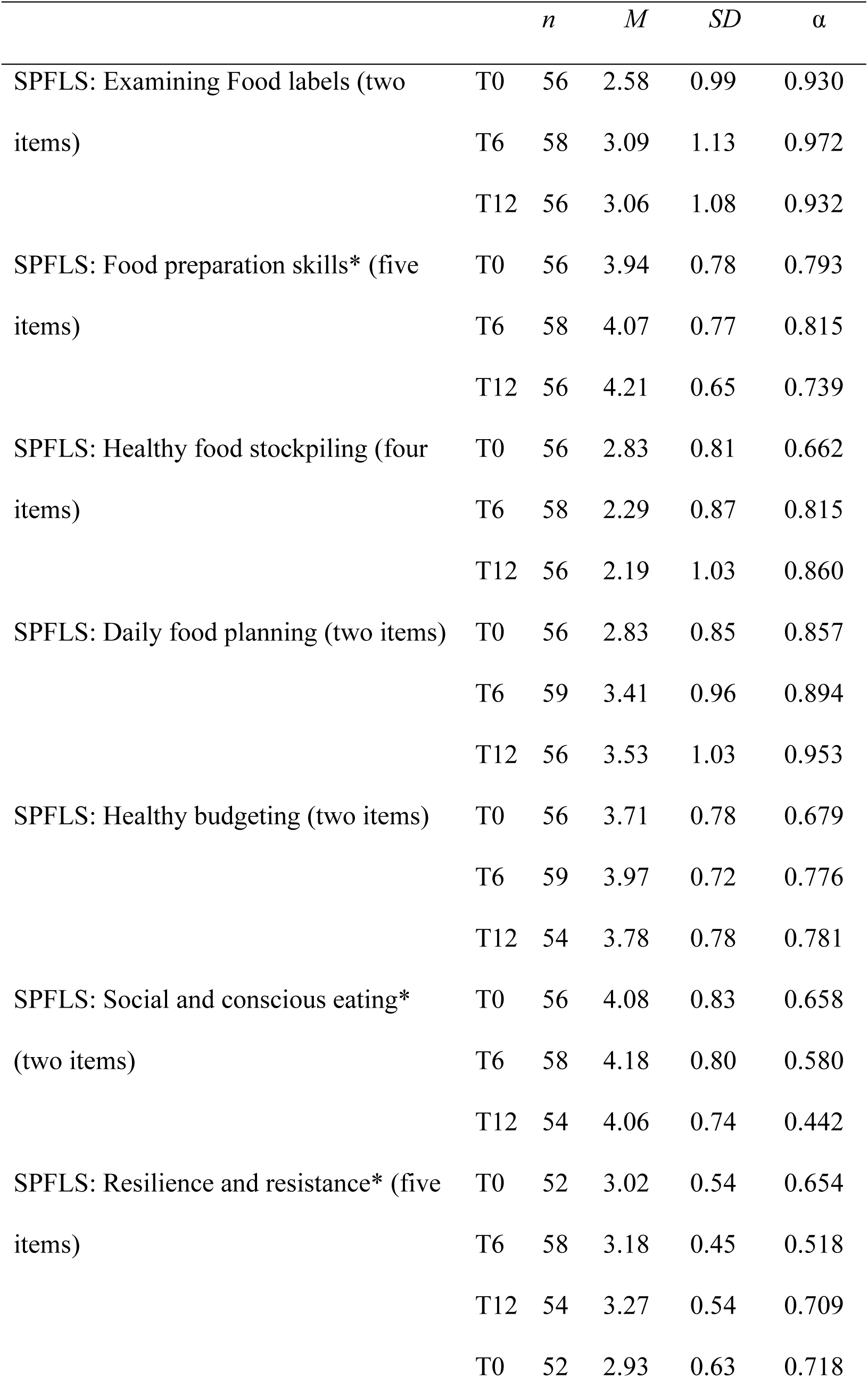
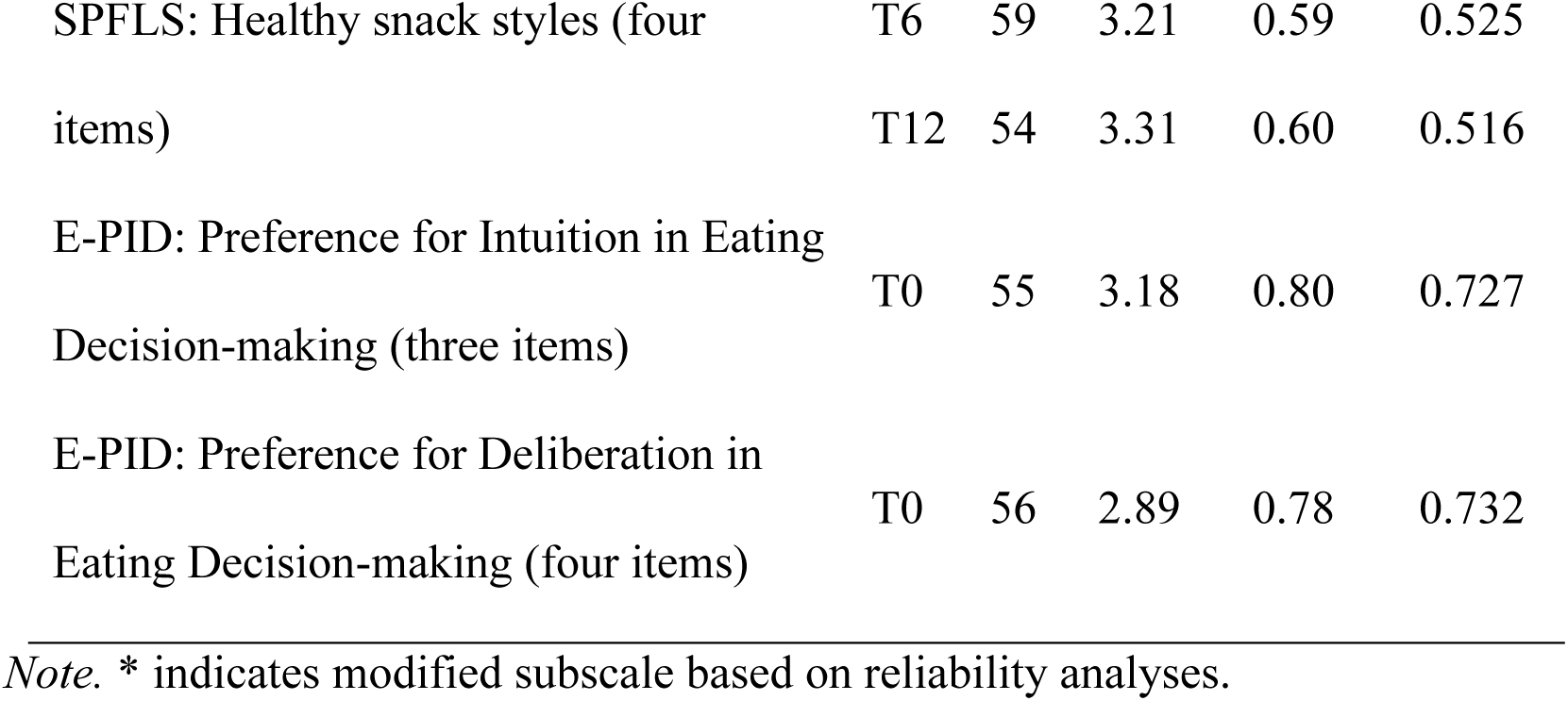
Means (M), standard deviations (SD) and internal consistencies (Cronbach’s a) of SPFLS and EPID subscales.

Height and weight were measured at the GP practices at T0, T6 and T12. BMI was calculated as kg/m².

#### 2.2.2. Effect modifiers and control measures

Preference for Intuition and Deliberation in Eating Decision-making (E-PID) was assessed on a seven-item scale [21, 29, 30]. The E-PID distinguishes between intuitive decision-making, which relies on gut feelings and immediate reactions, and deliberative decision-making, which involves careful consideration and analysis of information before arriving at a choice, through the respective sub-scales. Responses were assessed at T0 on five-point Likert scales (1 = I do not agree; 5 = I agree), and the mean was calculated per subscale. The subscales showed good internal consistencies (see Table 2).

Furthermore, screening for eating disorders was administered at T0 and T12 using the screening questions of the Patient Health Questionnaire (PHQ) regarding bulimia nervosa and binge-eating disorder [31]. Additionally, a question about night eating syndrome [32], which is not included in the PHQ, was incorporated. Responses were classified as eating disorder potentially present yes/ no.

### 2.4. Statistical analyses

Only participants completing all three assessments were included in the respective analyses; they did not significantly differ from participants who dropped out in sociodemographic characteristics or the number of previous weight-loss attempts [17]. Yet, there were missing values on variables relevant to the analyses, ranging from 2% for the consumption frequencies at T6 to 24% for night-time eating at T12. The number of included participants per analysis therefore varies due to listwise deletion.

The analyses were performed using IBM SPSS version 29. Descriptive data are presented as means (M) and standard deviations (SD). Changes in food intake (HEI) and food literacy (SPFLS total score and subscales) over 12 months were analyzed using one-way analyses of variance (ANOVA) followed by post-hoc Bonferroni comparisons. Relationships between weight loss, quality of food intake (via HEI) and food literacy (SPFLS total score) were analyzed using Pearson correlations. To investigate whether the change in HEI and SPLFS varied based on E-PID, one-way analyses of covariance (ANCOVA) was conducted with the E-PID subscales as covariates. Changes in the consumption frequency of individual food items were assessed using Friedman tests, due to the ordinal nature of the outcome data. To evaluate changes in eating disorder symptoms, which were assessed as categorical variables, McNemar tests were applied. For most analyses, *p* <.05 was considered statistically significant; for the SPLFS and its subscales, *p* <.006 was used to correct for multiple testing.

## 3. Results

### 3.1. Sample

Of 98 participants enrolled in study by June 2023, 61 (62%) completed the 12-month HAPpEN program (see Figure 1 for the participant flow). Their mean age at baseline was 47.6 ± 11.9 years, and 69.6 % of participants were women (*n* = 39). Their mean weight at baseline was 118.6 ± 22.6 kg and the body mass index (BMI) at baseline averaged 40.3 ± 6.5.

**Figure 1.**
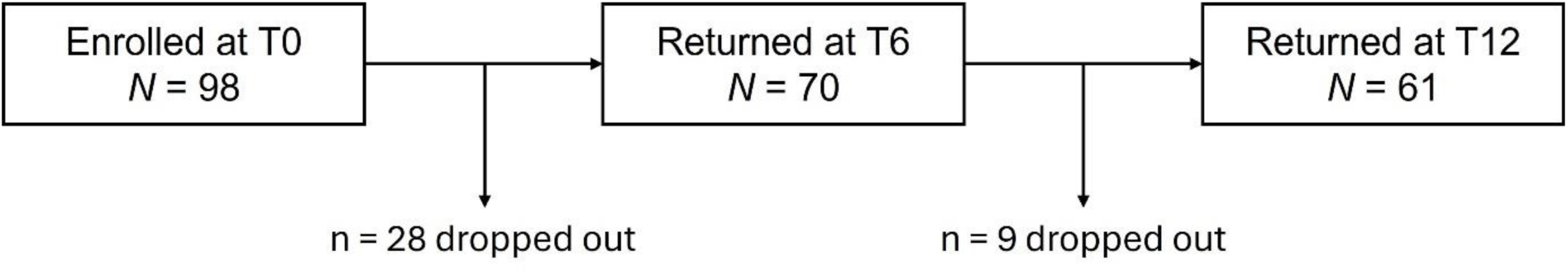
Participant flow chart. Of the *N* = 98 participants enrolling at baseline, *n* = 61 completed the assessment at T12.

Participants, on average, reported a somewhat higher preference for intuition (3.18 ± 0.80) versus deliberation (2.89 ± 0.78).

### 3.2. Does the intervention lead to changes in food intake after six and 12 months?

The mean HEI at baseline was 11.49 ± 3.72, reflecting an unfavorable dietary pattern. After six months, food intake showed a significant improvement, resulting in an HEI of 15.43 ± 3.61, which indicates a normal dietary pattern (*p* <.001). This enhancement remained statistically significant at T12, with an HEI of 14.79 ± 3.78, although there was no further statistically significant change between T6 and T12 (see Table 3).

**Table 3.**
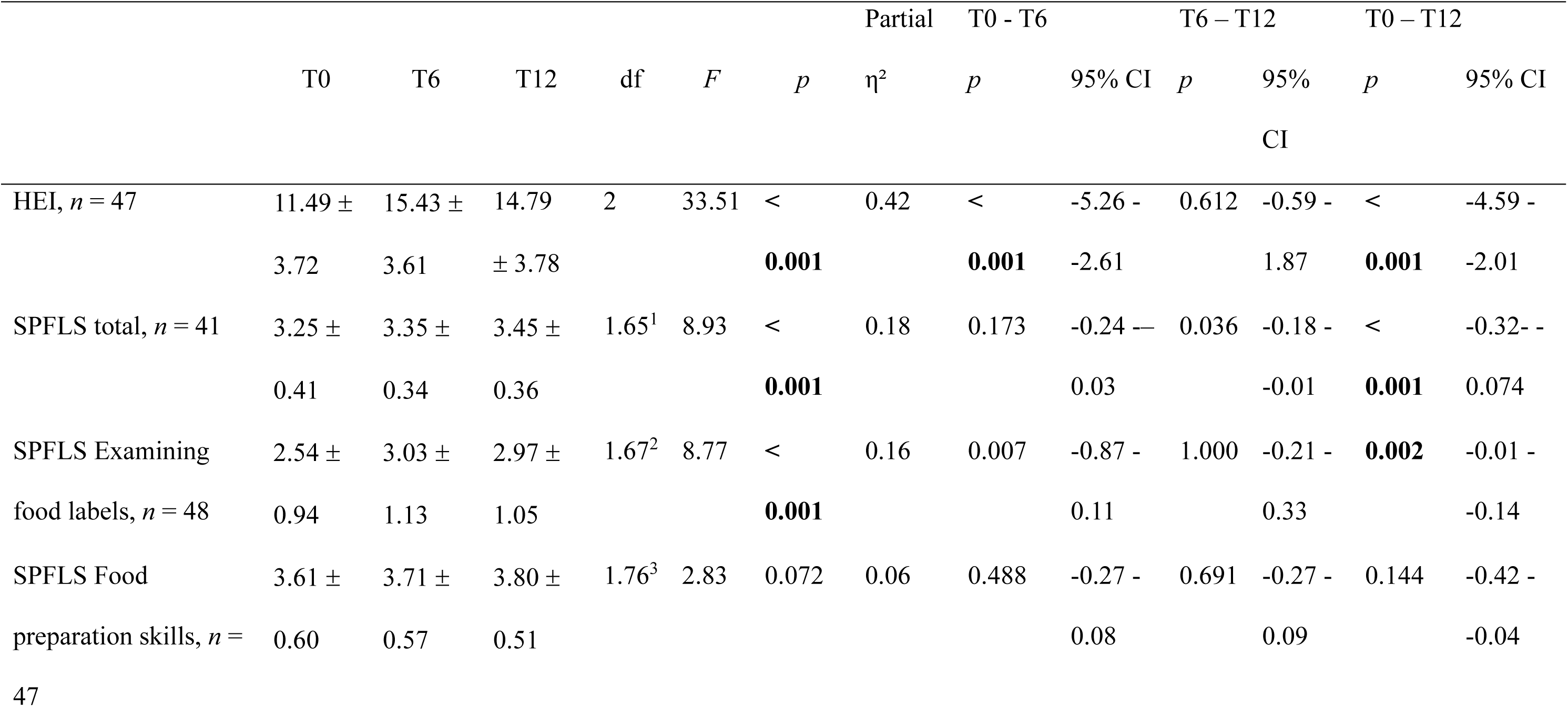
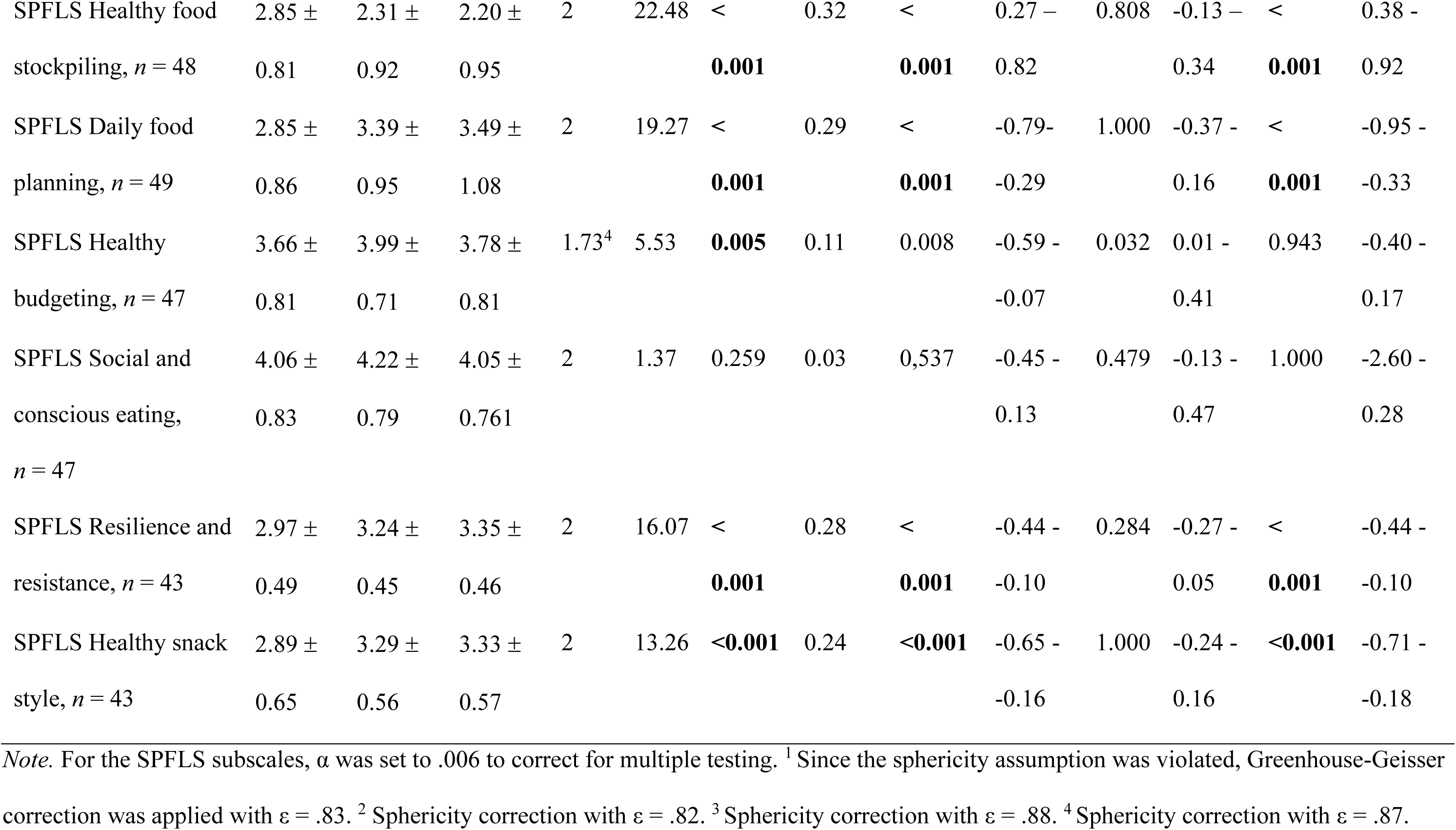
Mean values of HEI and SPFLS and results of the ANOVA to test for changes over time (T0, T6, T12). Statistically significant results are arked in bold.

When analyzing the individual items of the FFQ (see Table 4), a significant reduction was observed in the consumption frequency of highly processed foods, snacks and sausage and cold cuts. In contrast, the consumption frequency of fruits, raw vegetables, and pulses increased. There was no change for cooked vegetables, meat, fish, poultry, and eggs.

**Table 4.** Descriptive analysis of single items from the food frequency list with 6 = almost daily, 5 = several times per week, 4 = about once a week, = several times per month, 2 = once a month or less, 1 = never. Statistical analyses were performed using Friedman’s test. Statistically significant.

|  | T0 | T6 | T12 | p | X<sup>2</sup> | df | n N |
| --- | --- | --- | --- | --- | --- | --- | --- |
| **Fast Food** |  |  |  |  |  |  |  |
| Hamburger, doner | 2.21 ± 0.69 | 1.85 ± 0.57 | 1.87 ± 0.74 | < **0.001** | 17.54 | 2 | 53 |
| Fried- and currywurst | 2.58 ± 0.72 | 2.10 ± 0.69 | 2.13 ± 0.72 | < **0.001** | 21.12 | 2 | 52 |
| Fish fingers | 2.08 ± 0.77 | 1.80 ± 0.63 | 1.73 ± 0.60 | **0.007** | 9.82 | 2 | 51 |
| Frozen pizza | 1.98 ± 0.94 | 1.79 ± 0.85 | 1.67 ± 0.71 | **0.039** | 6.47 | 2 | 52 |
| Frozen ready meals | 1.79 ± 0.92 | 1.58 ± 0.83 | 1.46 ± 0.64 | **0.009** | 9.42 | 2 | 52 |
| French fries | 2.49 ± 0.89 | 2.11 ± 0.80 | 2.08 ± 0.65 | **0.002** | 12.71 | 2 | 53 |
| **Snacks** |  |  |  |  |  |  |  |
| Other sweets (candy, compote) | 3.26 ± 1.50 | 2.51 ± 1.19 | 2.23 ± 1.25 | < **0.001** | 15.17 | 2 | 53 |
| Ice cream, pudding | 2.98 ± 1.08 | 2.44 ± 1.06 | 2.73 ± 0.99 | **0.001** | 13.40 | 2 | 52 |
| Cakes, pastries, biscuits | 4.36 ± 1.08 | 3.25 ± 1.27 | 3.42 ± 1.23 | < **0.001** | 48.71 | 2 | 53 |
| Chocolate, chocolates | 3.92 ± 1.37 | 2.75 ± 1.25 | 3.04 ± 1.36 | < **0.001** | 35.01 | 2 | 52 |
| Salted snacks such as salted peanuts, crisps, and others | 3.42 ± 1.23 | 2.55 ± 1.03 | 2.60 ± 1.18 | < **0.001** | 23.89 | 2 | 53 |
| **Animal products** |  |  |  |  |  |  |  |
| Meat (without sausages) | 4.60 ± 0.82 | 4.24 ± 0.90 | 4.51 ± 0.97 | 0.052 | 5.91 | 2 | 53 |
| Sausages, ham | 5.13 ± 0.96 | 4.53 ± 1.20 | 4.45 ± 1.37 | <**0.001** | 20.35 | 2 | 53 |
| Fish | 3.25 ± 1.05 | 3.42 ± 1.05 | 3.36 ± 0.98 | 0.289 | 2.49 | 2 | 53 |
| Poultry | 4.09 ± 0.97 | 4.15 ± 0.93 | 4.21 ± 1.03 | 0.808 | 0.43 | 2 | 53 |
| Eggs | 4.17 ± 1.00 | 3.96 ± 1.11 | 4.08 ± 1.05 | 0.156 | 3.71 | 2 | 53 |
| **Fruits / vegetables** |  |  |  |  |  |  |  |
| Salad or vegetable, raw | 4.64 ± 0.98 | 5.21 ± 0.95 | 5.21 ± 0.77 | <**0.001** | 27.70 | 2 | 53 |
| Vegetable, cooked | 4.33 ± 1.00 | 4.60 ± 1.03 | 4.48 ± 1.06 | 0.067 | 5.40 | 2 | 52 |
| Fresh fruits | 4.91 ± 1.06 | 5.36 ± 1.00 | 5.42 ± 0.89 | <**0.001** | 22.52 | 2 | 53 |
| Pulses | 2.90 ± 1.26 | 3.38 ± 1.12 | 3.27 ± 1.35 | **0.002** | 12.47 | 2 | 52 |

Regarding beverages, the frequency of consumption of sugary soft drinks, fruit juices, and beer decreased, whereas water was consumed more frequently.

### 3.3. Does the intervention lead to a change in food literacy after six and 12 months?

The SPFLS total score increased significantly from T0 to T12, rising from 3.25 ± 0.41 at baseline to 3.45 ± 0.36 at T12 (*p* <.001); between T0 and T6, there was no statistically significant change (T6: 3.35 ± 0.34; *p* =.612). Concerning the subscale scores, significant improvements were observed in five out of eight domains (*p*s <.001; examining food labels, daily food planning, healthy budgeting, resilience and resistance, healthy snacks). The domain “healthy food stockpiling” also changed significantly but decreased over time from 2.85 ± 0.81 at T0 to 2.20 ± 0.95 at T12 (*p* <.001). The subscales “social and conscious eating” and “food preparations skills” remained unchanged. Results are summarized in Table 3.

### 3.4. Control analysis: Are changes in food intake and food literacy related to changes in weight and BMI?

Food literacy as measured by the SPFLS total score and diet quality as measured by the HEI were positively correlated at T6 (*r* =.49, *p* <.001) and T12 (*r* =.42, *p* =.002), indicating medium to large effects [33]. HEI at T6 was negatively correlated with relative changes in BMI (*r* = -.32, *p* =.016) and weight (*r* = -.32, *p* = 0.016) from T0 to T6 with medium effects [33], reflecting that weight loss was greater in individuals with better HEI scores 6 months into the intervention. For food literacy at T6, correlations were also negative, but did not reach statistical significance (*r*s ≤ -.32, *p*s ≥.083). For relative changes in BMI and weight from T0 to T12 and from T6 to T12, associations with food literacy and intake were also negative and indicated small effects [33], but not statistically significant (see Table 5 for all correlations).

**Table 5.** Pearson correlations between food literacy (SPFLS total score), food intake (diet quality assessed via HEI) and relative change in BMI and weight between T0, T6 and T12.

|  | Food literacy (SPFLS<br>total score) |  |  | Food intake (diet quality<br>assessed via HEI) |  |  |
| --- | --- | --- | --- | --- | --- | --- |
|  | *r* | *p* | *n* | *r* | *p* | *n* |
| Food literacy and intake measured at T6 |  |  |  |  |  |  |
| Relative change in BMI<br>from T0 to T6 | -.23 | .083 | 56 | -.32 | .016 | 57 |
| Relative change in weight<br>from T0 to T6 | -.23 | .085 | 56 | -.32 | .016 | 57 |
| Food literacy and intake measured at T12 |  |  |  |  |  |  |
| Relative change in BMI<br>from T0 to T12 | -.23 | .095 | 54 | -.26 | .064 | 53 |
| Relative change in weight<br>from T0 to T12 | -.23 | .095 | 54 | -.26 | .064 | 53 |
| Relative change in BMI<br>from T6 to T12 | -.15 | .315 | 50 | -.09 | .526 | 48 |
| Relative change in weight<br>from T6 to T12 | -.15 | .315 | 50 | -.09 | .526 | 48 |

### 3.5. Does the change in food intake or food literacy differ depending on E-PID?

According to the ANCOVAs (see Table 6), most main effects of time on the outcomes were rendered nonsignificant once the covariate was included, with the exception of the SPLFS subscale “Healthy food stockpiling” when preference for deliberation was included as a covariate; healthy food stockpiling was thus affected by the intervention independent of the participants’ preference for deliberation. There was also only one statistically significant interaction effect between time and preference for intuition for the “food preparation skills” SPLFS subscale, suggesting potential differences in intervention effectiveness for this outcome depending on the patients’ preference for intuition. Furthermore, a preference for deliberation was significantly associated with the SPLFS subscales “daily food planning” and “examining food labels”.

**Table 6.**
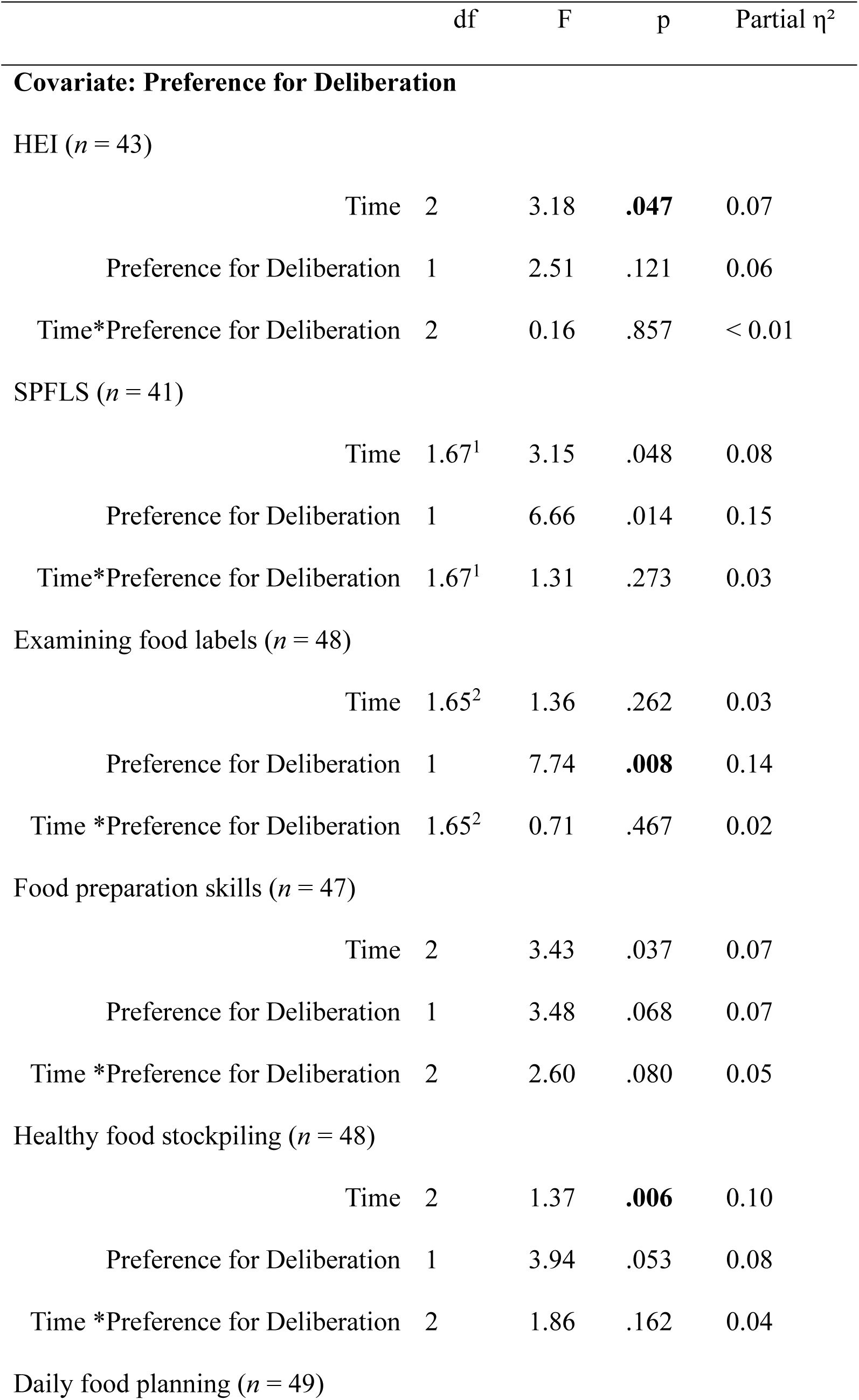
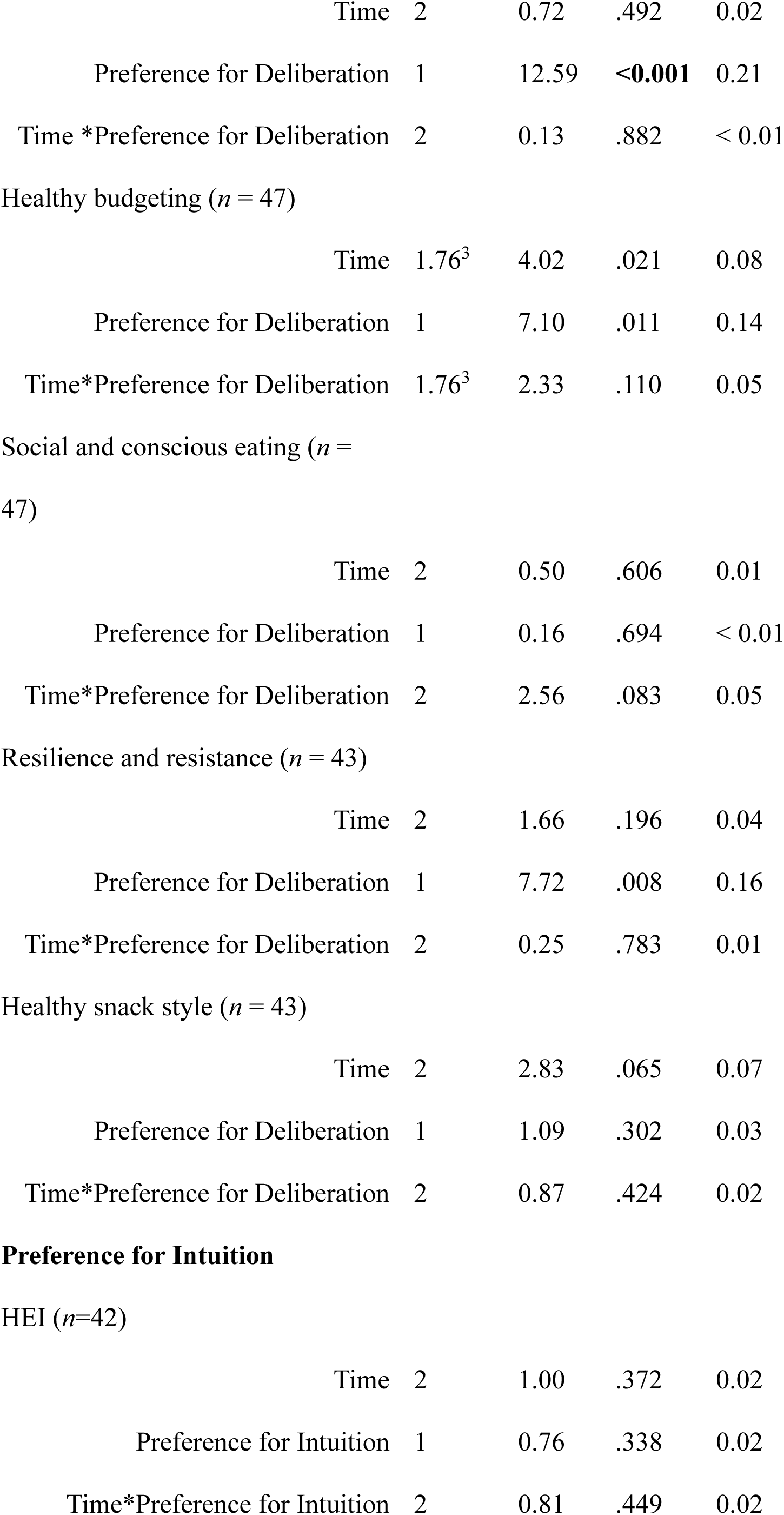
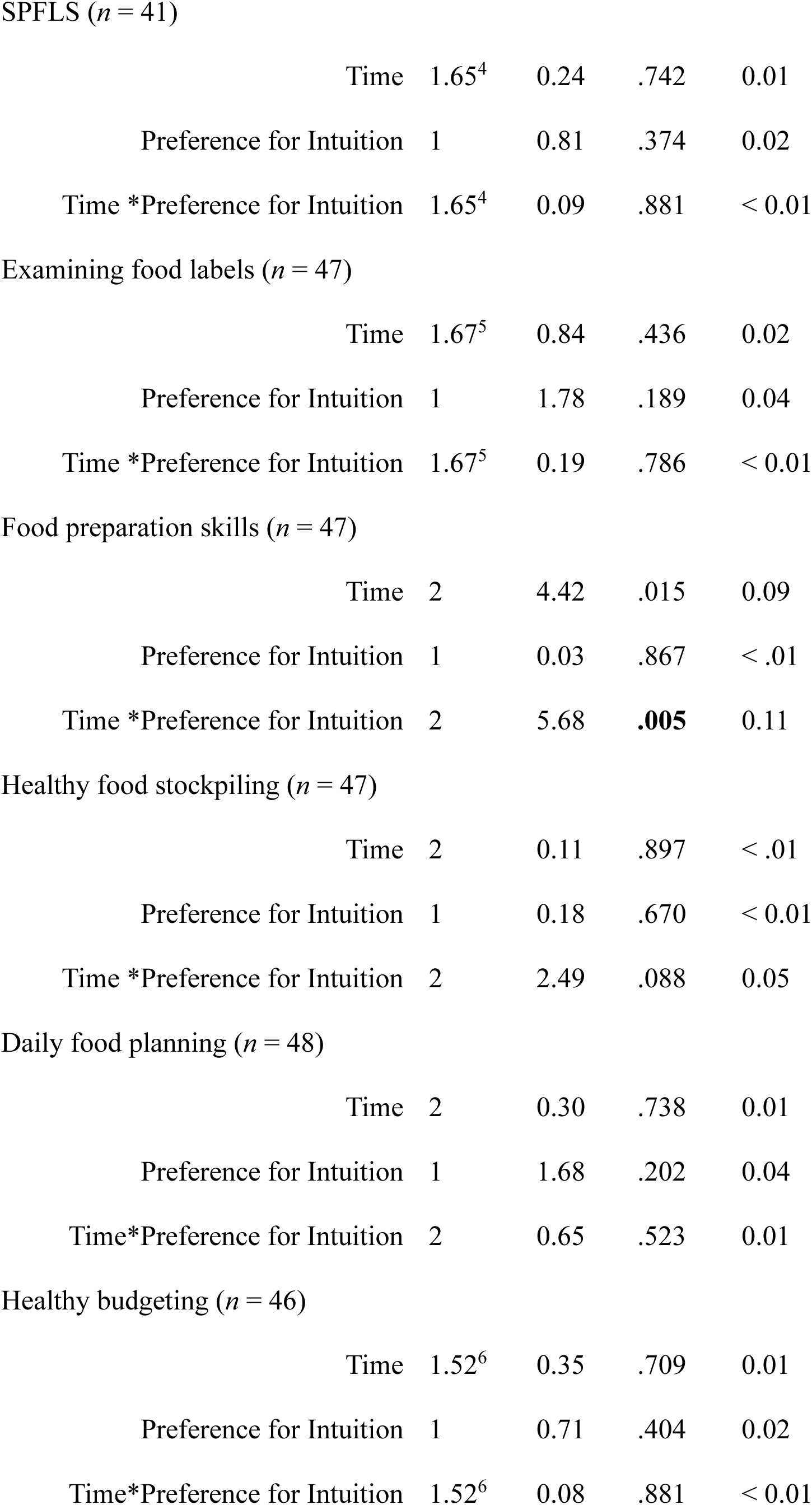
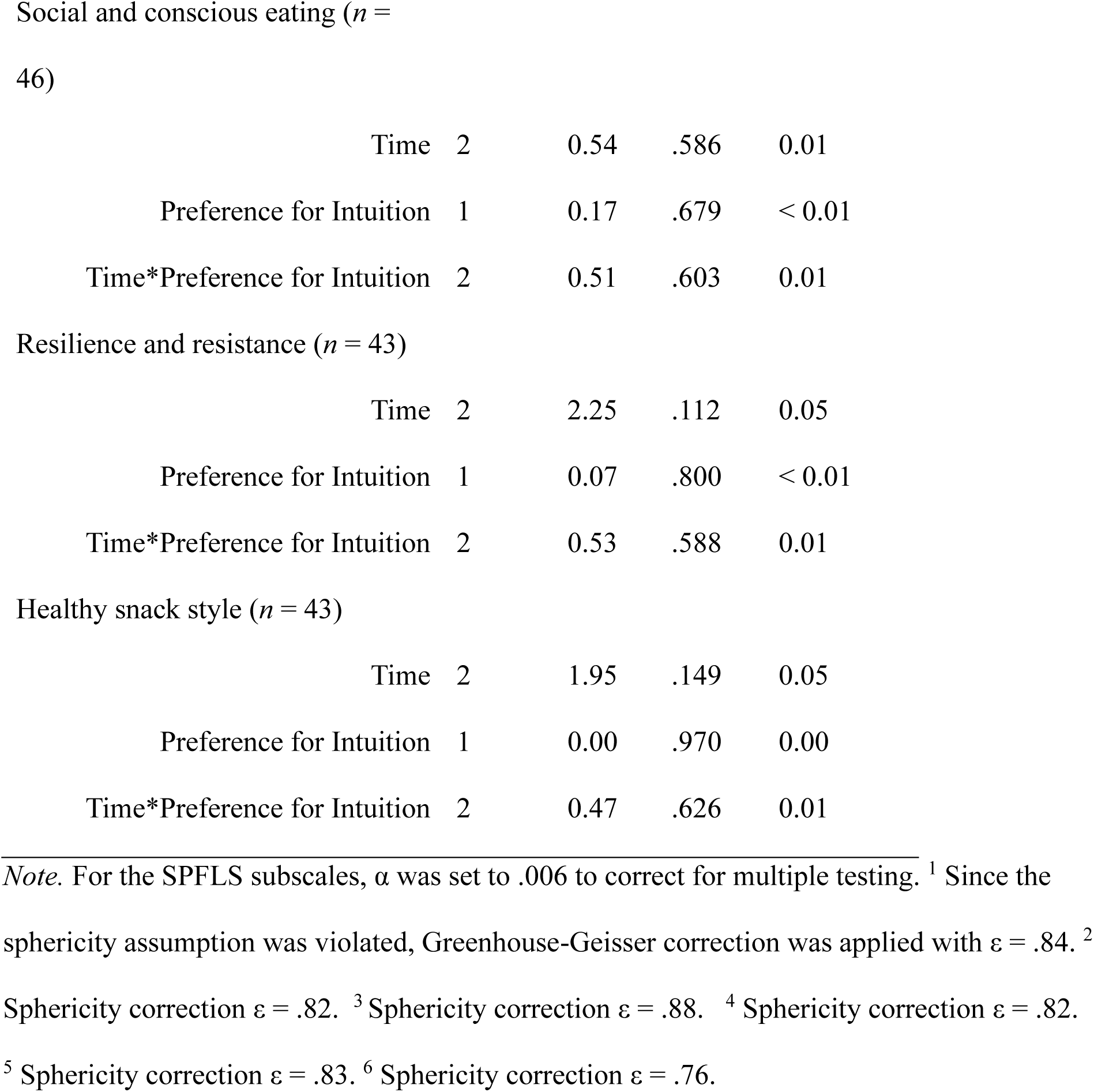
Results of the ANCOVA testing for modulating effects of E-PID. Statistically significant results are marked in bold.

### 3.6. Does the intervention result in changes to eating disorder symptoms?

Screening for bulimia nervosa, binge-eating disorder, and night-eating syndrome indicated that one participant may have suffered from bulimia nervosa at the start of the program, as well as a positive response to the night-eating syndrome question in twelve individuals. No participant suffered from binge-eating disorder at either time point. There was no statistically significant change over the course of the program, indicating that the program did not lessen, but also nor worsen, eating disorder symptoms (see Table 7).

**Table 7.** Descriptive and statistical analysis of PHQ screening question eating disorders. No cases of binge eating were detected. For statistical analysis, two-sided McNemar tests were.

|  | T0 | T12 | *p* | *n* |
| --- | --- | --- | --- | --- |
| Bulimia nervosa | 1.8 % (*n* = 1) | 3.6 % (*n* = 2) | 1.000 | 51 |
| Binge-eating disorder | - | - | - | - |
| Night eating syndrome | 23.1 % (*n* = 12) | 8,2 % (*n* = 4) | .180 | 42 |

## 4. Discussion

The HAPpEN pragmatic trial tested a multicomponent GP-led and digitally enhanced intervention for obesity management. In line with the reduction in body weight and metabolic risk factors [17], the present analysis indicates that participants significantly improved their food literacy and intake, and that these improvements were one potential underlying reason for weight loss. The intervention furthermore did not increase eating disorder symptoms, despite putting self-monitoring and goal setting at its core, providing further evidence for (digital) self-monitoring interventions being low-risk approaches to weight management [22]. However, potential interindividual differences based on personality traits such as the preferred decision-making style require further attention.

Mirroring the results on weight loss [17], also food intake improved considerably in the first six months of program participation and then stabilized without further statistically significant changes. This is in line with the energy balance model of obesity [18] which indicates that weight loss results from a negative energy balance which can be achieved through consuming fewer calories, e.g. via consuming fewer energy-dense foods [23]. The HAPpEN intervention successfully achieved this goal, as was not only indicated by a general improvement of eating patterns, but also by a specific reduction in the consumption frequency of fast foods, sweet and savory snacks, sugar-sweetened and alcoholic beverages, as well as an increase in the consumption frequency of salads, raw vegetables, fruits, and water. This further underlines the potential of breaking the complex behavior change process down into smaller, more manageable steps to increase motivation and perceived ability to change [34] by (1) providing guidance on the consumption of specific foods or food groups and by tailoring (2) the choice of target foods to the needs of the individual and (3) the goals to current levels of knowledge and behavior.

Interestingly, from a mechanistic perspective, food literacy seemed to have changed significantly only after changes in dietary behavior became apparent, which was indicated by a significant change in the SPFLS total score only occurring after one year, but not after six months of program participation. However, changes occurred already after six months for several subscales, including “daily food planning” and “healthy snack style”. Indeed, these were aspects directly targeted by the goal setting component of the intervention. Interestingly, participants also seemed to have felt more empowered to resist unhealthy foods, as indicated by changes in the subscale “resilience and resistance”. Other subscales showed no statistically significant change, including “food preparation skills” and “social and conscious eating”, however, these were also not directly targeted by the intervention. Some group-based activities were offered, such as a joint cooking event, but these were offered on a voluntary basis and rarely, which may not have been sufficient to induce changes at the group level.

Even more importantly, the majority of these events took place after the T6 assessment, which may explain the smaller and sometimes non-significant changes between baseline and T6 compared to T12. Since prior research demonstrated positive effects of culinary interventions on weight loss outcomes [35], these should be considered as a more central component in future iterations of the program to further boost food literacy.

The present results furthermore support the notion that a preference for deliberation in eating decision-making may be somewhat beneficial for healthy eating [29], given its associations with the ability to plan and read food labels. Furthermore, when controlling for the decision-making style preferences, most main effects on food intake and food literacy were no longer statistically significant. While this may hint at decision-making style preferences influencing whether an intervention is effective, as proposed in prior research [21], the results have to be interpreted with caution given the small sample size. Replications are warranted in larger samples to test whether an intervention relying on self-regulatory behavior change techniques such as self-monitoring and goal setting are indeed more appealing to and more effective in individuals with a preference for deliberation [21]. Further research is also needed to uncover how individuals with different decision-making style preferences interact with specific intervention components to test whether they differ in engagement with certain features of multicomponent interventions to allow for tailoring.

The pragmatic nature of the HAPpEN trial allowed testing the program’s effectiveness in a real-life setting. However, several limitations arose from this context which need to be acknowledged. Most importantly, a control group only provided weight measurements but did not complete questionnaires, so that changes in food intake and literacy could only be analyzed over time in the intervention group. Changes in literacy and behavior therefore could also be attributed to social desirability or measurement reactivity [36] instead of to the intervention. The questionnaire only assessed food consumption frequency, so potential changes in portion sizes could not be investigated. Further, the sample size was small, which precluded the reliable detection of smaller effects. Finally, we only tested one specific intervention package and engagement in the various components was highly difficult to assess. The present pragmatic evaluation thus does not provide insights into the relative importance of intervention components. In-depth evaluations, e.g. via factorial trials [37], are necessary to explore which intervention components were most helpful in inducing changes in food literacy and intake to reduce intervention complexity and resulting burden in the future.

The present research demonstrates that a GP-led, digitally supported obesity management program can successfully improve food literacy and promote healthier diets in rural populations that translate into initial weight loss and then weight maintenance. By integrating a stepped approach for tailored goal setting in a multicomponent blended intervention, applicability and success in a diverse sample of overweight adults living in a rural area could be achieved, providing a blueprint for further obesity prevention efforts in rural areas.

## Data Availability

All data produced in the present study are available upon reasonable request to the authors

## List of abbreviations

ANCOVA: Analysis of Covariance
ANOVA: Analysis of Variance
BMI: Body-Mass Index
E-PID: Preference for Intuition and Deliberation in Eating Decision-making
FFQ: Food Frequency Questionnaire
GP: General Practitioner
HAPpEN: Hausarzt-zentriertes Adipositas-Präventionsprogramm: Exercise & Nutrition
HEI: Healthy Eating Index
PHQ: Patient Health Questionnaire
SPFLS: Self-Perceived Food Literacy Scale

## Acknowledgement

We express our gratitude to our patients and GPs for their participation. Special thanks to Asarnusch Rashid, Nico Jordan, Madeleine Keßler (Centre for Telemedicine, Bad Kissingen), Susanne Tittlbach, Tina Bartelmeß, Constanze Betz, Alisa Bader, Miriam Witthüser, Bria-Estella Johnson (University of Bayreuth), and Antonia Baldauf (Friedrich-Alexander University Erlangen-Nuernberg) for their support and contributions.

## Sources of support

This work was supported by the Bavarian Federal Ministry of Health, Care and Prevention (G31i-G8000–2021/2630–23, 07.06.2022). The funder had no involvement in the study design; in the collection, analysis and interpretation of data; in the writing of the report; and in the decision to submit the article for publication.

## Author contributions

AW: Data curation, formal analysis, writing – original draft

MH: Conceptualization, funding acquisition, investigation, methodology, project administration, writing – review & editing

RH: Conceptualization, funding acquisition, project administration, writing – review & editing

MaM: Conceptualization, methodology, writing – review & editing

NvS: Conceptualization, funding acquisition, investigation, writing – review & editing

LMK: Conceptualization, formal analysis, methodology, supervision, visualization, supervision, writing – original draft

## Author declarations

The authors declare no conflicts of interest.

## Notes

### Competing Interest Statement

The authors have declared no competing interest.

### Clinical Trial

DRKS00033916

### Author Declarations

The HAPpEN trial has been approved by the University of Bayreuth ethics committee (Az. O 1305/1 - GB).

